# Circulating tumor DNA: An alternative to tissue biopsy for detecting EGFR mutation in NSCLC

**DOI:** 10.1101/2025.09.19.25336178

**Authors:** Md Kabirul Islam Soroar, Sharmistha Roy, Hosne Ara, Sharif al-Nur, Riyadh Arifin Akanda, Mohammad Masum Alam

## Abstract

**Background:** Non-small cell lung cancer (NSCLC) patients who have mutations in their EGFR gene respond more favorably to Tyrosine Kinase Inhibitors (TKIs) than to standard chemotherapy. However, tissue biopsy-based EGFR testing is invasive, costly, and technically challenging. Plasma-derived circulating tumor DNA (ctDNA) offers a minimally invasive and cost-efficient alternative for mutation profiling. This study assessed the agreement between EGFR mutation detection in plasma-derived ctDNA and tissue biopsy in NSCLC patients from tertiary care hospitals in Bangladesh.

**Methods:** In this cross-sectional analytical study, we recruited 32 patients with NSCLC before EGFR-TKI treatment. EGFR mutations in ctDNA samples were identified using the Amplification Refractory Mutation System (ARMS) PCR method. Tissue biopsy results were obtained from routine diagnostic procedures. Agreement between ctDNA and tissue biopsy results was assessed using kappa statistics, and diagnostic performance metrics were calculated.

**Results:** Most of our study participants were male (75%) and had stage IV lung adenocarcinoma (72%). We observed substantial agreement between plasma-derived ctDNA samples and tissue biopsies (kappa, κ = 0.683). This agreement was almost perfect (κ = 0.826) when calculated for patients with stage IV disease. The overall concordance was 84.4%. Compared with tissue biopsy, ctDNA testing yielded a sensitivity of 73.3% and specificity of 94.1%.

**Conclusion:** Plasma-derived ctDNA demonstrates substantial agreement with tissue biopsy for EGFR mutation detection in patients with NSCLC, particularly those with advanced-stage disease. These findings support ctDNA as a viable alternative for molecular profiling in settings where tissue biopsy is limited or impractical.

## INTRODUCTION

Cancer stands as a leading cause of mortality worldwide. In 2022, it was responsible for overall 16.8% of deaths and accounted for 22.8% of deaths related to non-communicable diseases. Among all cancer types, lung cancer stands out as the most lethal, giving rise to around 2.5 million incident cases and 1.8 million deaths each year (1). In Bangladesh, it is the third most frequently diagnosed cancer and the second leading cause of cancer mortality (2).

NSCLC is responsible for nearly 85% of all lung cancer (3). For accurate categorization and targeted therapy, mutation analysis of NSCLC is necessary and mandated by different societies (4–6). Among the various mutations identified in NSCLC, the Epidermal Growth Factor Receptor (EGFR) mutation exhibits the highest prevalence among Asian patients. It is detected in approximately 55% of cases in East Asia and between 23% and 44% of cases in India. However, their prevalence among Caucasian patients is notably lower, ranging from 10% to 15% (7–9). Notably, about 93% of EGFR mutations are detected within exons 18 to 21 (10).

NSCLC patients harboring mutations in the EGFR tyrosine kinase domain are eligible for targeted therapy with tyrosine kinase inhibitors (TKIs). TKIs radically overturned the conventional chemotherapy regimen by almost doubling the progression-free survival (8). Detecting EGFR mutations in tissue is considered a prerequisite for starting TKI treatment and predicting treatment responses and prognoses. For this purpose, the College of American Pathologists has recommended testing of the cell block (from excisional biopsy or small biopsy) or cytological sample (e.g. smear) with adequate cellularity and at least 20% or more malignant cell content (4). However, lung cancer is often detected at an advanced stage, at which point excisional biopsy becomes impractical. Therefore, small biopsies and cytological specimens are the only sources for morphological assessment and mutation analysis (11). These biopsy procedures are invasive and can cause complications such as hemoptysis, pneumothorax and pneumonitis (12). Additionally, the results of EGFR mutation analysis can be influenced by the tumor sampling location, as a biopsy from one part of a tumor may not capture the complete molecular heterogeneity (13). Furthermore, pathological evaluation reveals malignant cells in approximately 73% of specimens (14), often necessitating repeat biopsies to get enough tissue for genetic testing.

The limitations of tissue biopsies can be overcome using a rapidly emerging technique, known as liquid biopsy. In this technique, the plasma or serum of a patient with cancer is used to separate circulating tumor DNA (ctDNA). It is then used for mutation analysis. ctDNA analysis offers multiple benefits over conventional tissue biopsy. First, sample collection for ctDNA is less invasive. Therefore, ctDNA testing can serve as an early screening tool for NSCLC. Second, it can detect minimal residual disease after intervention and can be used to detect disease recurrence (15,16). Third, ctDNA has a half-life of approximately 114 min (17). Therefore, it can be used to track disease status and drug responsiveness in real time. Fourth, serial ctDNA analyses during treatment can detect resistant mutations, such as T790M (18).

In recent years, researchers have attempted to determine whether ctDNA can replace tissue biopsy or is complementary to it. Numerous studies have used ctDNA to predict EGFR-TKI treatment responses and assess progression-free survival as a primary outcome. Many of these studies have yielded promising results (15,19). Several studies have also assessed the concordance and agreement between ctDNA and tissue biopsies, primarily across Europe, the United States, and the Asia-Pacific region. However, research from South Asia remains limited, with no published data currently available from Bangladesh. Therefore, this study assessed the agreement between EGFR mutation profile in ctDNA and tissue biopsies in NSCLC patients attending tertiary care hospitals in Bangladesh.

## MATERIALS AND METHODS

### Study design and participants

This cross-sectional analytical study was conducted at Bangabandhu Sheikh Mujib Medical University, now known as Bangladesh Medical University, in Bangladesh, over a one-year period from March 2024 to February 2025. Ethical clearance for this study was granted by the university’s Institutional Review Board. The sample size was calculated using the sample size calculation method for the agreement test of a two-rater study on a dichotomous variable (20). Considering the availability of patients and the duration of the study, a total of 32 patients with lung cancer were recruited using a consecutive sampling approach from the oncology inpatient and outpatient departments of several tertiary care hospitals across the country. Participants were aged ≥ 40 years and had histologically confirmed NSCLC. The eligibility criteria included no prior treatment with systemic anti-cancer drugs, including TKIs, and the availability of an EGFR mutation test result from tissue biopsy within one week of blood sample collection. After obtaining informed written consent from the study participants, an initial evaluation was performed by history-taking. The clinical history of the participants and EGFR test results from tissue biopsy were subsequently recorded in a pre-formed data collection sheet.

### cfDNA extraction and EGFR mutation analysis

Whole blood was collected in two four mL K_**2**_EDTA tubes for ctDNA extraction from each study participant. Following collection, the samples were processed within two hours. Initially, they were centrifuged at 3,000 × g for 10 minutes to separate plasma. Subsequently, a microcentrifugation at 11,000 × g for 10 minutes was performed to produce clear samples for mutation analysis. The samples were stored at −80 °C. ctDNA extraction and mutations analysis were conducted at the Department of Biochemistry and Molecular Biology of the University. The ctDNA was extracted using the CatchGene Catch-cfDNA Serum/Plasma 1000 Kit (CatchGene Co., Ltd., Taiwan).

EGFR mutation status was assessed using the AmoyDx EGFR 29 Mutations Detection Kit (Amoy Diagnostics, Xiamen, China) on a Rotor-Gene Q (72 wells) real-time PCR machine (Qiagen, Hilden, Germany). This kit is an amplification refractory mutation system (ARMS)-based PCR kit which qualitatively detects 29 somatic mutations in the EGFR gene, especially targeting exons 18 (G719X), 19 (deletions), 20 (exon 20 ins9, T790M, and S768I), and 21 (L858R and L861Q). Each PCR run included seven test samples, one positive control, and one no-template control, and was subjected to 46 cycles of amplification. PCR data were interpreted in accordance with the manufacturer’s guidelines.

### Statistical analysis

Data from this study were analyzed using the Statistical Package for Social Sciences (ver. 27) software. Chi-square and Fisher’s exact tests were applied to compare the EGFR mutation status across various clinicopathological characteristics. Agreement between plasma-derived ctDNA samples and tissue biopsies were assessed using the kappa statistics. Additionally, the diagnostic performance of plasma samples was assessed in comparison with that of tissue biopsies. Statistical significance was assigned to results with p-values ≤ 0.05.

## RESULTS

### Clinicopathological characteristics of study participants

This study included a total of 32 NSCLC patients. Key clinicopathological parameters of the participants are summarized in Table 1. The mean age of the participants was 59.2 (59.2 ± 10.9) years. Most of them were male (n=24, 75%) and had adenocarcinoma histology (n=30, 94%). The TNM staging of the tumor was determined using the International Association for the Study of Lung Cancer’s 9^th^ edition of the lung cancer TNM staging system. Among the study participants, 23 (72%) had stage IV disease.

**Table 1:** Clinicopathological characteristics and EGFR mutation frequency of the study subjects (n = 32).

| Clinicopathological characteristics | Frequency, n (%) |
| --- | --- |
| Overall | 32 (100%) |
| Age: 59.2 ± 10.9 <sup>a</sup> |  |
| ≤ 60 years | 18 (56%) |
| > 60 years | 14 (44%) |
| Sex: |  |
| Male | 24 (75%) |
| Female | 8 (25%) |
| Smoking history: |  |
| Never smoker | 16 (50%) |
| Current/Ex-smoker | 16 (50%) |
| Histological type |  |
| Adenocarcinoma | 30 (94%) |
| Squamous cell carcinoma | 2 (6%) |
| Histological grading |  |
| Moderately differentiated | 21 (66%) |
| Poorly differentiated | 11 (34%) |
| Tumor stage (TNM) |  |
| Stage II | 1 (3%) |
| Stage III | 8 (25%) |
| Stage IV | 23 (72%) |
| EGFR mutation subtypes in tissue: |  |
| Exon 19 deletion | 8 (25%) |
| L858R | 5 (16%) |
| Others | 2 (6%) |
| Wild type <sup>b</sup> | 17 (53%) |
| EGFR mutation subtypes in plasma: |  |
| Exon 19 deletion | 6 (19%) |
| L858R | 5 (16%) |
| Others | 1 (3%) |
| Wild type <sup>b</sup> | 20 (62%) |

SD, standard deviation; TNM, Tumor, Node, Metastasis
<sup>a</sup> Data expressed as mean ± SD, <sup>b</sup> EGFR mutations absent or not detected

### EGFR mutation status

Of 32 study subjects, EGFR mutations were present in 15 (47%) tissue biopsy samples and mutations were identified in 12 (38%) plasma-derived ctDNA samples, with exon 19 deletion being the most frequent alteration in both specimen types (Table 1). One plasma mutation was a TKI-resistant mutation (T790M), whereas two of the tissue mutations were TKI-resistant (T790M and exon 20 ins9). Table 2 outlines the relationship between EGFR mutation status and various clinicopathological parameters of the patients. In plasma, EGFR mutation rates were significantly higher in patients aged > 60 years and in never smokers (p-values of 0.043 and 0.028, respectively). However, sex distribution, smoking history, histological grade, and tumor stage did not differ significantly across the groups. In tissue samples, mutations were significantly higher in never smokers (p-values of 0.013).

**Table 2:** Relationship between EGFR mutation status and clinicopathological characteristics (n = 32).

| Clinicopathological characteristics | Plasma EGFR status |  |  | Tissue EGFR status |  |  |
| --- | --- | --- | --- | --- | --- | --- |
|  | Mutated | Wild-type <sup>a</sup> | *p* | Mutated | Wild-type <sup>a</sup> | *p* |
| Overall | 12 | 20 |  | 15 | 17 |  |
| Age |  |  | **0.043** <sup>b</sup> |  |  | 0.305 <sup>b</sup> |
| ≤ 60 years | 4 | 14 |  | 7 | 11 |  |
| > 60 years | 8 | 6 |  | 8 | 6 |  |
| Gender |  |  | 0.433 <sup>c</sup> |  |  | 1.0 <sup>c</sup> |
| Male | 8 | 16 |  | 11 | 13 |  |
| Female | 4 | 4 |  | 4 | 4 |  |
| Smoking History |  |  | **0.028** <sup>b</sup> |  |  | **0.013** <sup>b</sup> |
| Never smoker | 9 | 7 |  | 11 | 5 |  |
| Current/Ex-smoker | 3 | 13 |  | 4 | 12 |  |
| Histological grading |  |  | 0.139 <sup>c</sup> |  |  | 0.108 <sup>b</sup> |
| Moderately differentiated | 10 | 11 |  | 12 | 9 |  |
| Poorly differentiated | 2 | 9 |  | 3 | 8 |  |
| Tumor Stage |  |  | 0.103 <sup>c</sup> |  |  | 1.0 <sup>c</sup> |
| Stage II-III | 1 | 8 |  | 4 | 5 |  |
| Stage IV | 11 | 12 |  | 11 | 12 |  |

<sup>a</sup> EGFR mutations were absent or not detected; <sup>b</sup> Chi-square test; <sup>c</sup> Fisher's exact test.

### Agreement on EGFR mutation status

Of the 32 samples, 27 were concordant (11 positive and 16 negative), yielding a concordance rate of 84.4% (Tables 3 and 4). Among the 15 mutations present in the tissue sample, 11 were also positive in the plasma, resulting in a sensitivity of 73.3%. The overall specificity was 94.1%, with a positive predictive value (PPV) of 91.7% and a negative predictive value (NPV) of 80%. Concordance and sensitivity increased to 91.3% and 90.9%, respectively, when calculated from patients with stage IV disease. We also assessed the agreement between these two sample types (Table 3). Overall, we found a kappa (κ) value of 0.683, indicating substantial agreement between the two sample types. κ increased to 0.826 and reached almost perfect agreement when calculated from participants with stage IV disease (21).

**Table 3:** Comparison of EGFR mutations between plasma-derived ctDNA and tissue biopsy samples.

| ctDNA | Tissue |  |  | Kappa value, κ (95% CI) |
| --- | --- | --- | --- | --- |
|  | Mutated | Wild-type <sup>1</sup> | Total |  |
| Overall (n=32) |  |  |  |  |
| Mutated | 11 | 1 | 12 | 0.683 (0.432-0.933) |
| Wild-type <sup>a</sup> | 4 | 16 | 20 |  |
| Total | 15 | 17 | 32 |  |
| Stage IV tumor patients (n = 23) |  |  |  |  |
| Mutated | 10 | 1 | 11 | 0.826 (0.595-1.00) |
| Wild-type <sup>a</sup> | 1 | 11 | 22 |  |
| Total | 11 | 12 | 23 |  |

<sup>a</sup> EGFR mutation absent or not detected

**Table 4:**
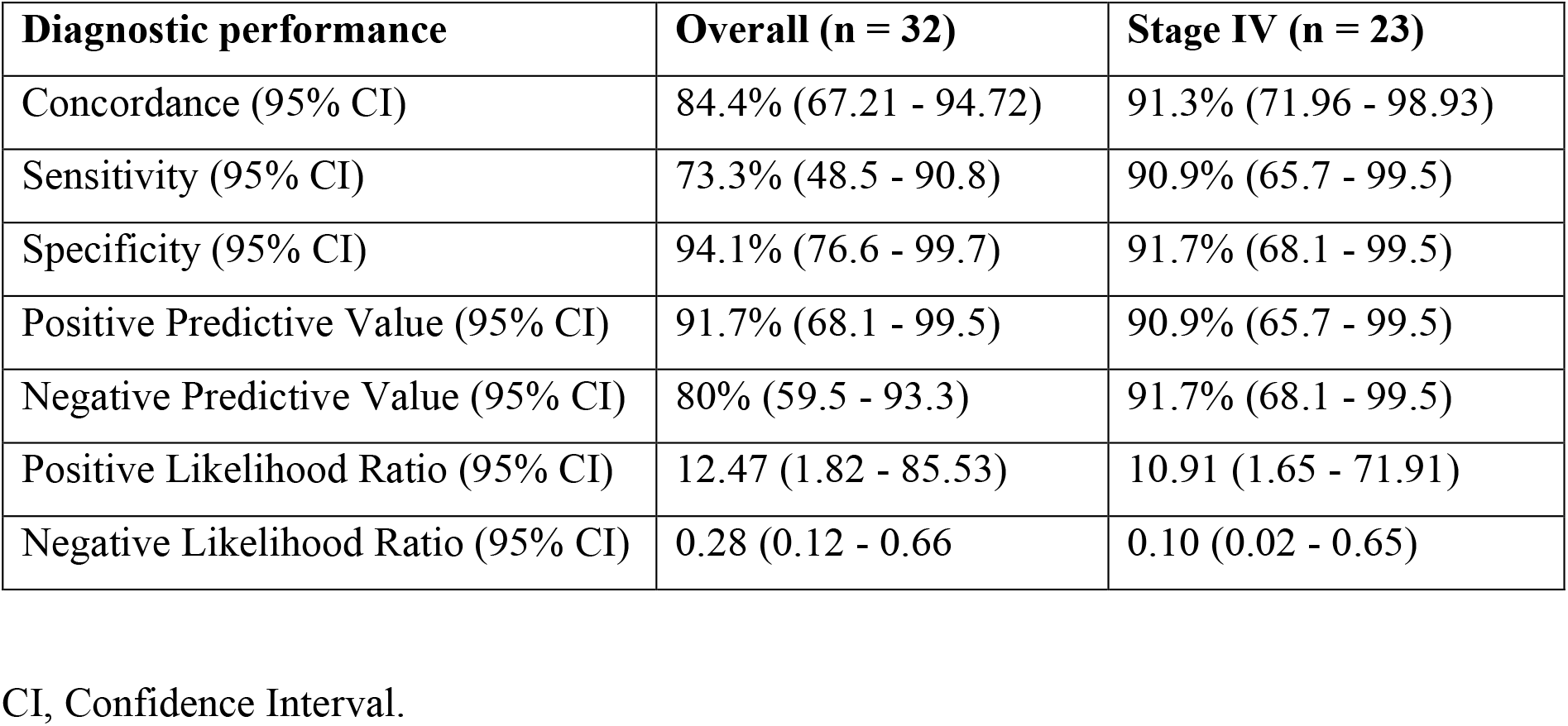
Diagnostic performance of plasma-derived ctDNA samples compared to tissue biopsy samples for detecting EGFR mutation in NSCLC.

## DISCUSSION

Based on the currently available data, no prior study from Bangladesh has compared EGFR mutation detection between tissue biopsies and ctDNA samples. Our study showed good agreement and concordance between these two.

The majority of the participants were aged 55 - 70 years, with a mean age of 59.2 years, consistent with previous studies (9,22). The PIONEER study, the most extensive epidemiological analysis in Asia, also found a similar mean age (23). Most of the subjects in our study were male (75%) and no statistically significant difference was found between sex distribution and mutation status in the plasma or tissue (*p-values* of 0.433 and 1.0, respectively). This result may reflect our relatively small sample size and imbalanced sex ratio (male: female = 3:1). Similar results were observed in some other studies (22,24–26). However, in the large database, female sex has been significantly associated with EGFR mutations, as in the IGNITE study (27).

Smoking is the most important factor for EGFR mutation detection, as EGFR mutations are more common in never smokers (23). In our study, half of the participants were never smokers (50%). However, 75% of the mutations in plasma (9 out of 12) and 73% of the mutations in tissue (11 out of 15) were detected in never smokers. In the IGNITE study, Han et al. (27) observed similar results. In their study, 67% of mutations in tissue samples and 64% of mutations in plasma samples were found in never smokers. In our study, 72% of the study participants had stage IV NSCLC. In ctDNA samples, we found that the EGFR mutation was strongly positive in this group (11 out of 23 stage IV subjects) compared to stages II–III. These findings were similar to those reported by Wulandari et al. (22). In the metastatic stage, the ctDNA content in the plasma sample of the patient becomes very high compared to that in the earlier stage, resulting in a high mutation detection rate in the metastatic stage (22).

In our study, we found good agreement and concordance between the tissue biopsy and ctDNA samples. We obtained an overall kappa value of 0.683, which increased to 0.826 when calculated from stage IV disease subjects. Our results were supported by some small- and large-scale studies (19,28,29). Soria-Comes et al. (28) reported an overall kappa value of 0.6, which increased to 0.7 when calculated from subjects with stage IV disease. Veldore et al. (29) published their study on 132 Indian patients having stage IV NSCLC with distal metastasis, where they used NGS to test plasma samples. Therefore, they found almost perfect agreement (κ = 0.931) between the plasma and tissue samples. In the IFUM study, we calculated a kappa value of 0.755 from the given data. In this study, the authors included patients with locally advanced or metastatic NSCLC, which was reflected in their high kappa values (19).

In our study, we also found a high concordance between tissue and plasma samples. The overall concordance rate was 84.4%. Compared with tissue biopsy, ctDNA demonstrated a sensitivity of 73.3%, specificity of 94.1%, PPV of 91.7%, and NPV of 80%. Concordance and sensitivity substantially increased when calculated from subjects with stage IV disease (concordance, 84.4% to 91.3%; sensitivity, 73.3% to 90.9%). Our study was also supported by some studies conducted in India, the Asia-Pacific region, and Europe (9,19,24,28,29). Three studies reported similar results in the Indian population. Joshi et al. (24) and Prabhash et al. (9) reported concordance of 83% and 82.9% respectively. Veldore et al. (29) did their study on stage IV disease subjects and found high concordance (98.48%), high sensitivity (94.44%) and perfect specificity (100%), which was very close to our results from stage IV disease. Soria-Comes et al. (28) published a study on the Spanish population, which provided results similar to those of our study (concordance, 87.4%; sensitivity, 70.6%; and specificity, 91.7%). The large IFUM study on the Caucasian population also reported similar results (concordance, 94.3%; sensitivity, 65.7%; specificity, 99.8%; and PPV, 98.6%) (19). Other studies from Asia have also reported similar concordance (ranging from 80% to 90%) (26,27,30,31). The ASSESS study (31), the IGNITE study (27) and Duan et al. (30) reported similar specificity, PPV and NPV. However, their published sensitivities were lower than those in our study (sensitivity ranged from 46% to 50%). This was because we had very few false-negative results. In our study, 11 out of 15 tissue EGFR-positive patients had stage IV disease, and from these 11 cases, we found that 10 of them also became positive in plasma. Therefore, we only had one false-negative result in this group. As a result, we observed a high overall sensitivity.

Therefore, this study indicates a good agreement between the EGFR mutations detected in the ctDNA samples and tissue biopsies. It was also found that the concordance, sensitivity, and specificity of ctDNA samples were very high compared to tissue biopsies, which is comparable to the findings in other parts of the world.

### Conclusion

We observed substantial agreement, which increased to almost perfect agreement in patients with Stage IV disease. This high level of agreement, along with high concordance, sensitivity, and specificity, strongly indicates that EGFR mutations can be reliably identified using ctDNA. These promising findings highlight the effectiveness of plasma-derived ctDNA as an alternative to tissue biopsy for detecting EGFR mutations in NSCLC patients in Bangladesh, especially when tissue biopsy is not feasible due to financial constraints or advanced disease stages, or when the tissue sample is inadequate for analysis.

### Limitation

This study had a limited sample size, with the majority of patients being male, resulting in a non-homogeneous distribution of the study population. In our study, EGFR mutation detection in tissue biopsies was performed as part of routine clinical investigations, and we performed it in ctDNA samples. This may have introduced operator-related variability.

### Recommendation

It is recommended that EGFR mutations be detected in tissue biopsies and ctDNA samples in the same center, possibly by the same operator, to minimize operator-related variability in the results. Future research should include a more balanced number of male and female patients to avoid any gender bias.

### Ethics Statement

Ethical approval for this study was granted by the Institutional Review Board of Bangabandhu Sheikh Mujib Medical University (Registration No. 5112, Date 13.07.2024). All procedures were conducted in accordance with the ethical standards outlined in the Declaration of Helsinki and relevant national guidelines. Patient data were anonymized prior to analysis to ensure confidentiality.

### Informed consent statement

Written informed consent was obtained from all study participants using a preformatted consent form, following a detailed explanation of the study’s purpose and procedures.

## Author Contributions

- **Study conception and design:** Md Kabirul Islam Soroar, Mohammad Masum Alam
- **Data acquisition and laboratory work:** Md Kabirul Islam Soroar, Sharmistha Roy, Hosne Ara
- **Statistical analysis and data interpretation:** Md Kabirul Islam Soroar, Sharmistha Roy, Sharif al-Nur, Riyadh Arifin Akanda, Mohammad Masum Alam
- **Study supervision:** Mohammad Masum Alam
- **Manuscript drafting, critical revision for important intellectual content, and final approval of the version to be published:** Md Kabirul Islam Soroar, Sharmistha Roy, Hosne Ara, Sharif al-Nur, Riyadh Arifin Akanda, Mohammad Masum Alam

All authors have read and approved the final manuscript and agree to be accountable for all aspects of the work, ensuring its accuracy and integrity.

## Conflict of Interest

**Md Kabirul Islam Soroar:** None declared.

**Sharmistha Roy:** None declared.

**Hosne Ara:** None declared.

**Sharif al-Nur:** None declared.

**Riyadh Arifin Akanda:** None declared.

**Mohammad Masum Alam:** None declared.

## Funding Statement

This research was fully funded by Md Kabirul Islam Soroar. Following its completion, he received a student thesis grant from Bangabandhu Sheikh Mujib Medical University. However, this grant had no role in the study design, data collection, statistical analysis, manuscript preparation, or the decision to publish. No other external funding, payment, or services from a third party were received by the authors or their institutions for any part of this work.

## Data availability statement

The datasets analyzed during the current study are available from the corresponding author upon reasonable request.

## Notes

### Competing Interest Statement

The authors have declared no competing interest.

### Author Declarations

Institutional Review Board of Bangabandhu Sheikh Mujib Medical University granted ethical approval for this work.

